# The Interplay Between Molecular Architecture, Pharmacology, and Suspected Adverse Drug Reactions Associated with Non-Steroidal Androgen Antagonists in The United Kingdom

**DOI:** 10.1101/2024.07.09.24309949

**Authors:** Simrit Dhillon, Albert A. Antolin, Alan M. Jones

## Abstract

**Aims:** To correlate potential links between the suspected adverse drug reaction (ADR) profile of licensed non-steroidal androgen receptor antagonists (NSARA) with their unique chemical properties and known off-target polypharmacology.

**Methods:** Physiochemical and polypharmacology data was curated from the Electronic Medicines Compendium, FDA New Drug Applications documents, and ChEMBL databases. System organ class (SOC, MedDRA) suspected ADRs and fatalities were curated from the United Kingdom Medicines and Healthcare products Regulatory Authority (MHRA) Yellow card spontaneous reporting scheme for their respective prescribing period; apalutamide (Jan 2019-), bicalutamide (Aug 2018-), enzalutamide (Aug 2018-), flutamide (Aug 2018-) and darolutamide (March 2019-) until Oct 2023. The number of daily doses (*dd*) was extracted from OpenPrescribing and NHS Digital secondary care medicines data. Data was standardised before comparison to suspected ADRs and fatality reports per 100,000 *dd*.

**Results:** A total of *n* = 2,480 suspected ADRs were associated with 42,903,000 *dd* of NSARAs in the United Kingdom. The highest number of ADRs were associated with enzalutamide (*n* = 1,091) and bicalutamide (*n* = 749). Enzalutamide was found to have the most off-target pharmacological interactions of the NSARAs studied (*n* = 5) including potent inhibition of γ-aminobutyric acid, GABA receptor (IC_50_ = 2.6 µM vs C_max_ = 7.7 µM) associated with nervous system disorders (*n* = 72, accounting for 73% of all NSARA ADRs in this SOC). Apalutamide, the only other GABA inhibitor (IC_50_ = 3 µM vs C_max_ = 2.9 µM) had the highest relative rate of suspected nervous system ADRs at 1.08 per 100,000 dd.

Apalutamide was also a modest inhibitor of the human Ether-à-go-go-Related Gene (hERG) ion channel (IC_50_ = 6 µM vs C_max_ = 2.9 µM) and had the highest rate of suspected cardiac arrhythmia ADRs, 30-fold over, enzalutamide, a significantly weaker hERG inhibitor (15.7 µM vs C_max_ = 7.7 µM).

Darolutamide was the only NSARA to show effects at 5-HT (serotonin) receptor at < 10 µM but did not translate to psychiatric disorders due to low clinical BBB penetration but a an association with hepatobiliary and cardiac disorders was identified based on this inhibitory axis. Suspected skin and subcutaneous SOC ADRs was associated with all NSARAs (except flutamide) but did not reach statistical significance (*P* = .25). A rationale for epidermis reactions relating to apalutamide containing a masked arylamine was explored but molecular matched pair (MMP) analysis with enzalutamide suggests it may not be a chemical cause. Statistical significance (*P* < .05) was identified in reported fatalities associated with NSARAs, flutamide had *n* = 24 or 897.5 fatalities per 100,000 *dd* which was likely due to both the indication and the small number of *dd* (*n* = 3,000) during the time period of the study.

**Conclusions:** An investigation of suspected ADRs, standardised to the number of *dd* for the novel NSARA drug class identified SOCs of potential interest. The highest number of reports related to enzalutamide and bicalutamide. Suspected skin and subcutaneous ADRs approached statistical significance and was interrogated for chemical and pharmacological connections for the first time with the aid of MMP analysis. A potential correlation to nervous system disorders and cardiac arrhythmia for the GABA and hERG inhibitors, enzalutamide and apalutamide, respectively was identified. Darolutamide’s interaction with 5-HT may influence ADRs associated with cardiac and hepatobiliary SOCs. Statistically significant number of suspected fatalities with flutamide was identified.

## Introduction

In the United Kingdom, 52,300 men per year are diagnosed with prostate cancer, [1] making it the most common cancer with peak diagnosis between 70-74 years old. [1] Hormone therapy is recommended as the first-line treatment strategy by the National Institute for Health and Care Excellence (NICE). [2-3] Androgen receptor antagonists (ARA) inhibit the exertion of androgen-like testosterone and dihydrotestosterone (DHT) pharmacological effects by competitively inhibiting testosterone binding to ARs [4]. Steroidal ARAs treat a range of conditions including sexual deviation and hot flushes with gonadorelin analogue therapy. [5]

Non-steroidal ARAs (NSARAs) are specifically used for the treatment of prostate cancer. NSARAs target prostate cancer cells by inhibiting androgen production, therefore, decreasing prostate cancer cell growth. [6] NSARAs can be either first generation (bicalutamide, flutamide) which exclusively target AR translocation to the nucleus or second generation (enzalutamide, apalutamide, darolutamide) which have further improvements on this mechanism of action (MoA). [7-8] Monitoring adverse drug reactions (ADRs) reported to the MHRA Yellow Card Scheme in the United Kingdom [9] is important as 1 in 16 patients admitted to hospital are suspected to be experiencing an ADR [10]. The ability to determine correlation of NSARAs ADRs to molecular structure and/or off-target pharmacological factors is therefore timely. [11-18]

## Methods

The licensed first generation NSARAs are bicalutamide [19] and flutamide [20]; and second generation, enzalutamide [21], apalutamide [22], darolutamide [23] (**Figure 1**) were selected based on inclusion and exclusion criteria of this study (**Table 1**).

**Figure 1.**
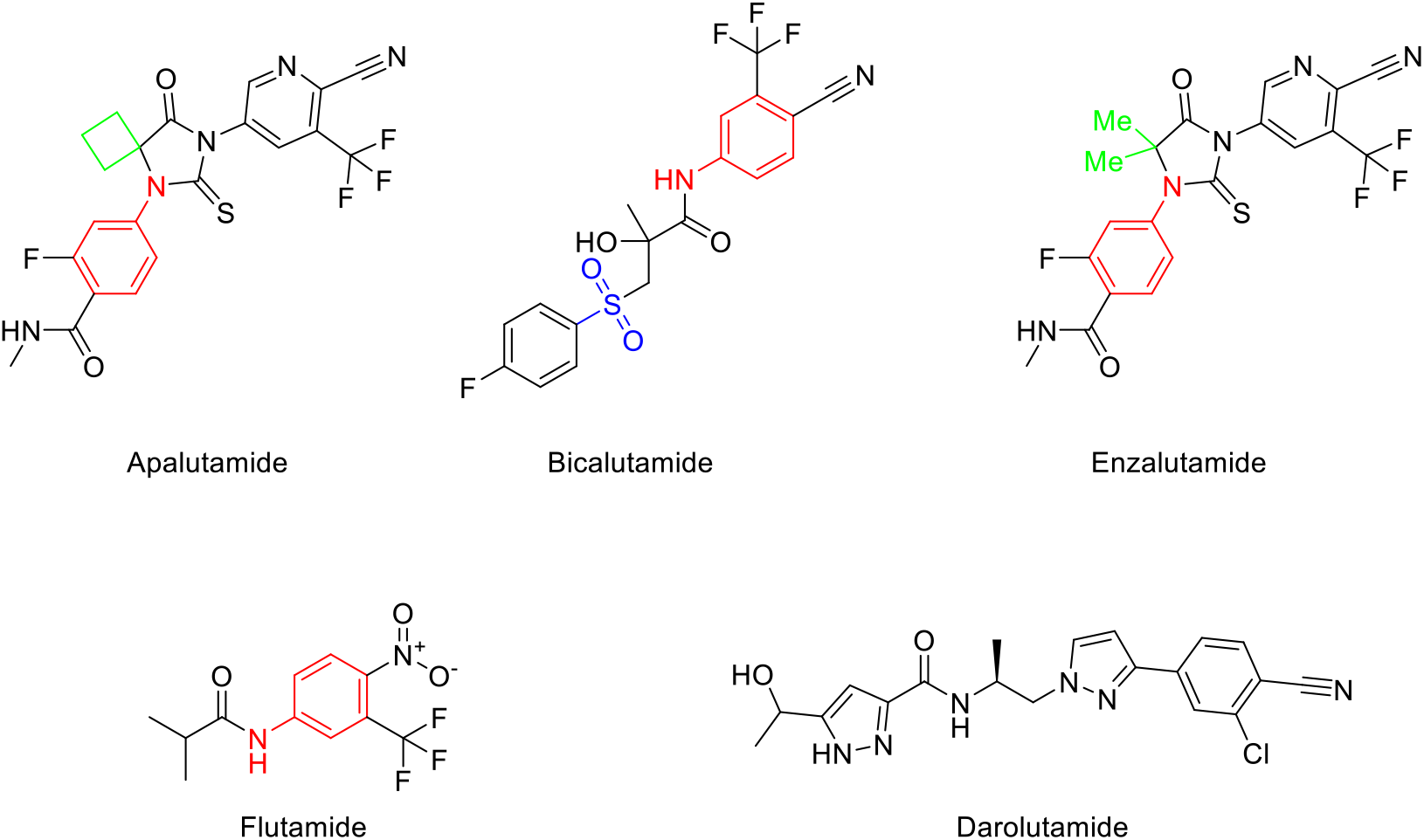
Structures of NSARAs studied. Colour coding: red for masked arylamine motif; blue for sulfonyl group; green for additional MMP analysis.

**Table 1.**
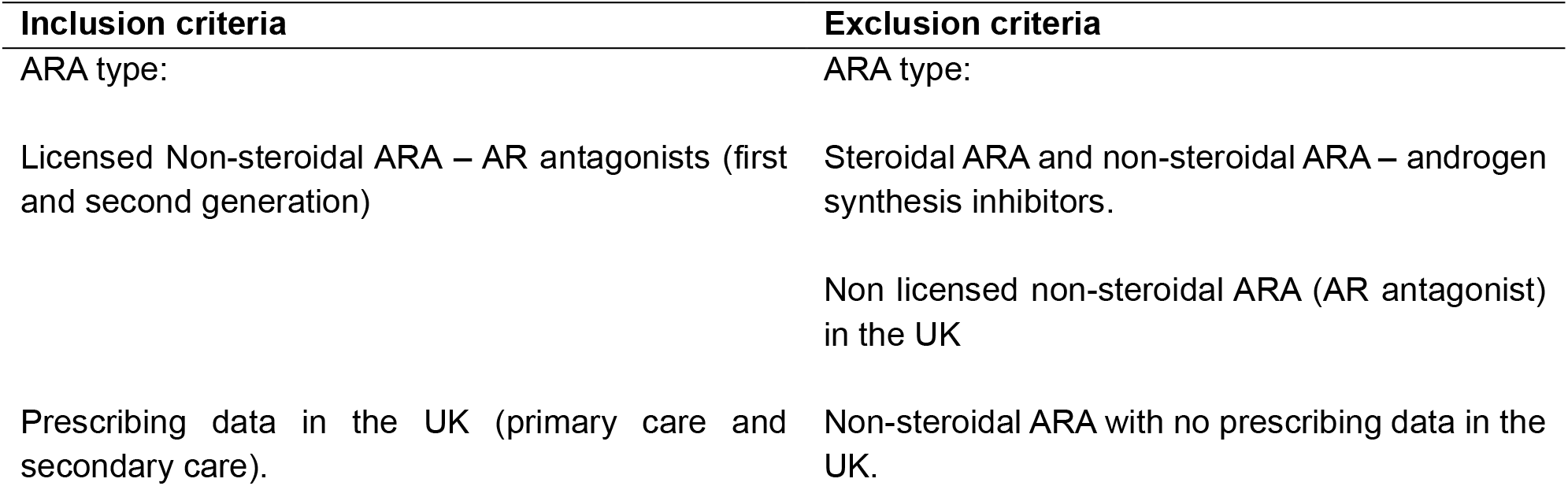
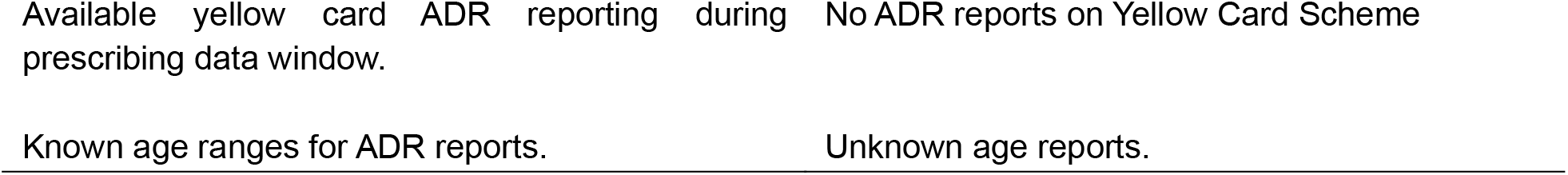
Inclusion and exclusion criteria.

| Inclusion criteria | Exclusion criteria |
| --- | --- |
| ARA type: | ARA type: |
| Licensed Non-steroidal ARA – AR antagonists (first and second generation) | Steroidal ARA and non-steroidal ARA – androgen synthesis inhibitors. |
|  | Non licensed non-steroidal ARA (AR antagonist) in the UK |
| Prescribing data in the UK (primary care and secondary care). | Non-steroidal ARA with no prescribing data in the UK. |

### Prescribing Data

NSARA data prescribing information was curated from OpenPrescribing database (for primary care prescribing [24]) and NHS Digital Secondary Care Medicines Data. [25] These datasets providied the tablet quantities of the NSARAs dispensed from each NHS trust.

### Drug pharmacology

NSARA’s molecular and physiochemical properties were extracted from the Chemical database of bioactive molecules with drug-like properties, European Molecular Biology Laboratory (ChEMBL), [26] FDA New Drug Application (NDA) documents, [27-30] and the Electronics Medicines Compendium (EMC). [31] Dosing regimens were extracted from the British National Formulary (BNF). [32]

The following parameters were calculated: pIC_50_ was calculated from the negative log of the average AR IC_50_ data. Lipophilic ligand efficiency (LLE) was calculated using the following equation: pIC_50_ – log_10_P. [33]. The predicted blood-brain barrier (BBB) penetration requirements were interrogated as follows: molecular weight (MW) <450 Da, <6 hydrogen bond donors (HBD), <2 hydrogen bond acceptor (HBA), a neutral or basic drug (indicated by pK_a_), topological polar surface area (tPSA) <90Å, log_10_D (at pH 7.4) between 1 to 3 and a low p-glycoprotein affinity. The C_max_ was converted from ng/mL to nM based on the unique molecular weight of each NSARA.

### Adverse drug reaction reporting

Suspected adverse drug reactions (ADRs) of all United Kingdom licensed NSARAs were extracted from the MHRA Yellow Card interactive drug analysis profile (iDAPs). [9] Suspected ADRs reported for apalutamide (since Jan 2019), bicalutamide (since Aug 2018), enzalutamide (since Aug 2018), flutamide (since Aug 2018), and darolutamide (since Mar 2019) with data extracted corresponding to the longitudinal timeline of each NSARAs prescribing in the United Kingdom.

Standardised NSARA ADRs were determined by calculating the ADR incidence rate per 100,000 daily doses (*dd*) for each drug:

1. Total tablets quantity dispensed divided by the respective standard of NSARA daily number of tablets = total *dd*.
2. Scale factor: Total *dd* dispensed / 100,000.
3. ADRs multiplied by the scale factor gave the ADRs per 100,000 *dd* of the NSARAs.

### Target Affinity

ChEMBL was used to extract the target proteins for each NSARA (accessed 29_th_ November 2023). [26] Where > 1 IC_50_ for a human protein target was available, the mean IC_50_ value was calcualated.

### Ethical Approval

Ethical approval was not required by the School of Pharmacy sub-ethics committee due to the use of fully anonymised patient data.

### Statistical Analysis

Excel for Microsoft 365 was used to perform chi-squared (χ_2_) tests on the standardised ADR’s/100,000 *dd* data. A *P* < .05 was set for statistical significance-.

## Results

### Physicochemical properties and pharmacokinetics

Properties of the NSARAs (clog_10_ P, pIC_50_, LLE) are shown in **Table 2**. Enzalutamide was the most lipophilic drug (clog_10_P = 3.99) and darolutamide was the most potent AR antagonist (pIC_50_ = 7.59). All AR antagonists had an LLE < 5. NSARAs that comply with the BBB criteria: bicalutamide, flutamide and darolutamide successfully passed the MW requirements; with apalutamide being the heaviest drug (477 Da). All the NSARA’s failed the guidelines for number of HBAs by having >2 HBA’s, however, passed the HBD criteria (<6). Apalutamide, enzalutamide and darolutamide are weak bases and bicalutamide and flutamide are neutral. Apalutamide, enzalutamide and flutamide had a _*t*_PSA < 90 Å. Bicalutamide and darolutamide met the criteria for log_10_D^7.4^ (2.71 and 2.44, respectively). Enzalutamide and flutamide were not P-glycoprotein substrates. Overall, flutamide passed the most BBB requirements out of all AR antagonists (achieving 5/7 BBB requirements), whilst apalutamide met the least (3/7 conditions).

**Table 2.**
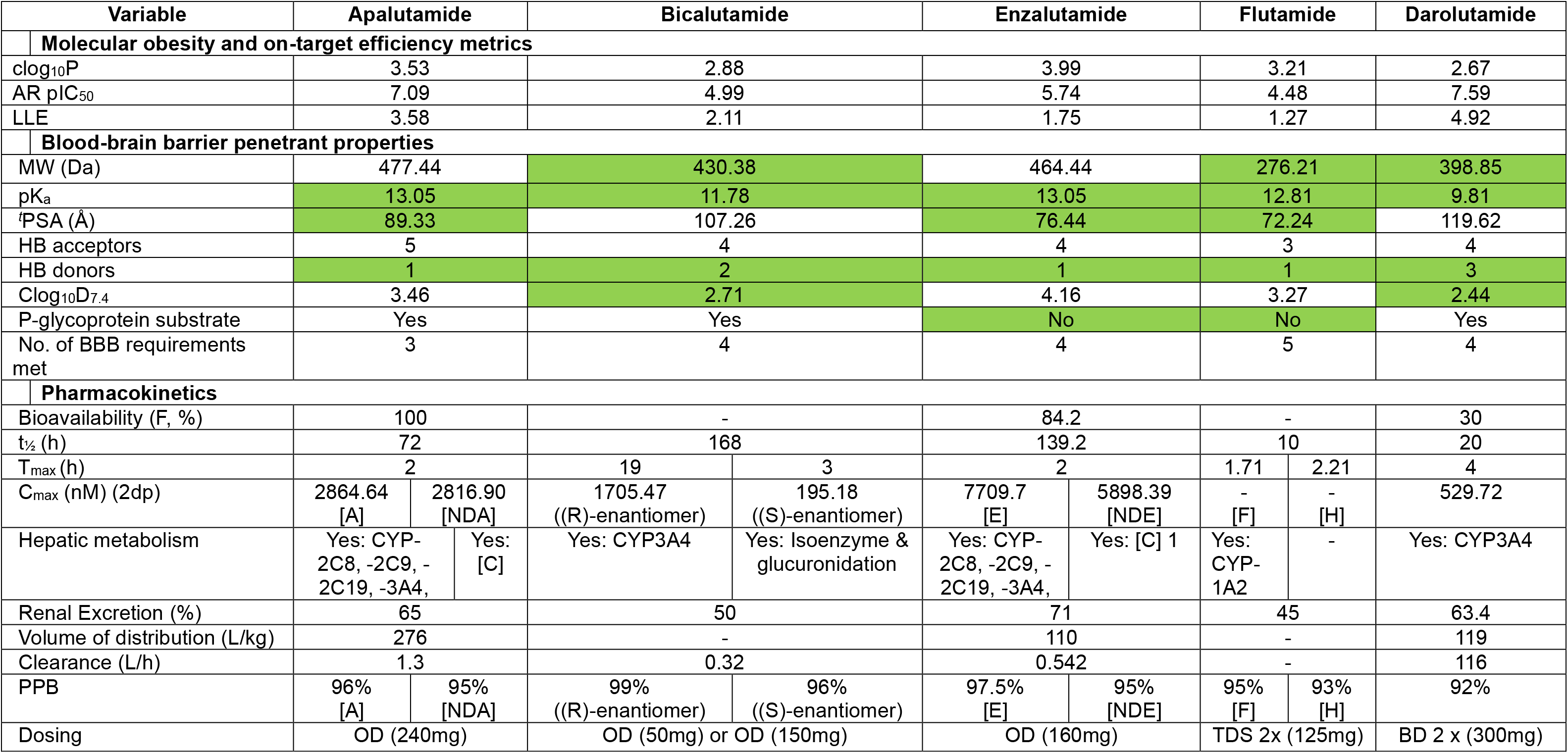
Summary of the physicochemical, blood-brain barrier predicted penetration, and pharmacological properties of the five NSARAs. Colour coded green met the BBB thresholds. Abbreviations: [A], apalutamide; [NDA], *N*-desmethyl apalutamide; [E], enzalutamide; [NDE], *N*-desmethyl enzalutamide; [F], flutamide; [H], hydroxyflutamide; [C], carboxylesterases ;clog_10_P, calculated by log_10_P; LLE, lipophilic ligand efficiency; MW, molecular weight; pKa, acid dissociation constant; ^*t*^PSA, total polar surface area; HB, hydrogen bond; clog_10_D^7.4^; calculated log_10_D at pH 7.4; BBB, blood-brain barrier; C_max_, peak serum concentration; T_max_, time taken to reach C_max_; PPB, plasma protein binding.

In clinical settings, darolutamide did not significantly alter cerebral blood flow (CBF), consistent with its low clinical BBB penetration and subsequent low risk of CNS-related adverse events. [34] However, a significant reduction in CBF was observed with enzalutamide suggestive of increased BBB penetration. [34] This was further supported by cerebrospinal fluid (CSF) for enzalutamide in rats showing a 3-6% penetration. [35] CSF measurement in dogs for apalutamide show a 2-5% penetration, [36] A single case report for CSF leak with flutamide was identified [37] but no clinical data was identified for bicalutamide.

Flutamide required the most daily tablets; with the regime consisting of two tablets, three times a day due to the short half-life (10 h). Apalutamide had the largest volume of distribution (276 L/kg) with bicalutamide and flutamide having undetermined volumes of distribution (V_d_).

### Target affinity

**Table 3** shows the strength of inhibition assessed by using the mean IC_50_ values. Apalutamide and darolutamide hadthe most potent inhibition of the AR receptors, with IC_50_ = 63 nM and 26 nM, respectively. Bicalutamide and flutamide had the weakest AR antagonist inhibition (1.2 and 1.3 µM, respectively). Bicalutamide had additional interactions with progesterone receptors at clinically achievable levels (IC_50_ = 1.8 µM vs C_max_ = 1.7 µM). Enzalutamide was found to have the most off-target pharmacological interactions of the NSARAs studied (*n* = 5) including potent inhibition of γ-aminobutyric acid, GABA receptor (IC_50_ = 2.6 µM vs C_max_ = 7.7 µM). Apalutamide, was the only other GABA inhibitor (IC_50_ = 3 µM vs C_max_ = 2.9 µM) and a modest inhibitor of the human Ether-à-go-go-Related Gene (hERG) ion channel (IC_50_ = 6 µM vs C_max_ = 2.9 µM). Enzalutamide was a significantly weaker hERG inhibitor (15.7 µM vs C_max_ = 7.7 µM). Furthermore, darolutamide was the only NSARA to show effects at 5-HT (serotonin) receptor at < 10 µM.

**Table 3.** Target pharmacology (mean IC_50_ (nM)) of the five NSARAs studied alongside C_max_ and clinical measurements for BBB penetration. n.r. = not reported.

|  | **Apalutamide** | **Bicalutamide** | **Enzalutamide** | **Flutamide** | **Darolutamide** |
| --- | --- | --- | --- | --- | --- |
| **AR** | **63** | **1167** | **806** | **1348** | **26** |
| **Glucocorticoid receptor** | - | - | <19500 | - | - |
| **Progesterone receptor** | - | 1819 | - | - | - |
| **Bile salts export pump** | - | 54450 | - | 79396 | - |
| **Progesterone receptor** | No inhibition | - | 16000 | - | - |
| **GABA** | 3000 | - | 2600 | - | - |
| **hERG** | 6000 | - | 15700 | - | - |
| **5-HT transporter** | - | - | - | - | <10000 |
| **C<sub>max</sub>** | **2865** | **1706** | **7710** | **n.r.** | **530** |
| **BBB penetration** | 2-5% (dog) [36] | n.r. | 3-6% (rat) [35] | CSF leak case report [37] | 0.3-0.6% (human) [34] |
|  | 10 – 100 nM |  |  |  |  |
|  | 100 – 1000 nM |  |  |  |  |
|  | 1000 – 10000 nM |  |  |  |  |
|  | > 10,000 nM |  |  |  |  |
| - | Undetermined |  |  |  |  |

### Total ADR’s and fatalities

ADR profiles of the NSARAs were standardised based on the number of *dd* dispensed in the time period of the study (**Table 4**). Overall, flutamide had the largest number of ADR’s/100,000 *dd* (15.8); followed by darolutamide (15.0) with enzalutamide the least (3.1). Flutamide had the largest number of fatalities/100,000 dd (*n* = 24 (897.5 per 100,000 *dd*)). Bicalutamide had the next highest at 0.4 deaths per 100,000 *dd*. Darolutamide was the only NSARA to have no reported deaths. For the selected SOC based on relative proportions of suspected ADRs and > 100,000 *dd* during the time period are shown in **Figure 2**.

**Table 4.** Summary of the suspected ADRs for the five NSARAs. Key: Total ADRs calculated by total ADRs / 100,000 dd; fatalities calculated by fatalities / 100,000 dd; p-values calculated by chi-squared test; *P* values exclude flutamide (apart from fatalities) and where there are n < 3 reports.

|  | **Apalutamide** | **Bicalutamide** | **Enzalutamide** | **Flutamide** | **Darolutamide** | **P** |
| --- | --- | --- | --- | --- | --- | --- |
| Number of dd /100,000 | 1700000<br>(17.0) | 5500000 (55.0) | 35500000 (355.0) | 3000 (0.03) | 200000 (2.0) | - |
| Total ADRs | 182 (10.93) | 749 (13.69) | 1091 (3.07) | 423 (15.82) | 35 (15.55) | .08 |
| Fatalities | 2 (0.12) | 22 (0.40) | 50 (0.14) | 24 (897.53) | 0 | < .05 |
| **Blood and lymphatic system disorders** |  |  |  |  |  |  |
| Total ADRs | 4 (0.24) | 3 (0.05) | 4 (0.01) | 0 | 1 (0.44) | .89 |
| Fatalities | 0 | 0 | 0 | 0 | 0 | - |
| Thrombocytopenia | 2 (0.12) | 0 | 1 (0.002) | 0 | 0 | - |
| Neutropenia | 0 | 0 | 1 (0.002) | 0 | 1 (0.44) | - |
| **Cardiac disorders** |  |  |  |  |  |  |
| Total ADRs | 13 (0.78) | 4 (0.07) | 12 (0.03) | 0 | 2 (0.89) | .70 |
| Fatalities | 1 (0.06) | 0 | 0 | 0 | 0 | - |
| Cardiac Arrhythmia's | 5 (0.30) | 0 | 4 (0.01) | 0 | 0 | - |
| Heart failure | 6 (0.36) | 0 | 0 | 0 | 1 (0.44) | - |
| **General disorders and administration site conditions** |  |  |  |  |  |  |
| Total ADRs | 34 (2.04) | 17 (0.31) | 121 (0.34) | 0 | 4 (1.78) | .52 |
| Fatalities | 1 (0.06) | 0 | 29 (0.08) | 0 | 0 | - |
| **Nervous system disorders** |  |  |  |  |  |  |
| Total ADRs | 18 (1.08) | 17 (0.31) | 72 (0.20) | 0 | 1 (0.44) | .82 |
| Fatalities | 0 | 0 | 0 | 0 | 0 | - |
| Neurological disorders NEC | 8 (0.48) | 8 (0.15) | 33 (0.09) | 0 | 1 (0.44) |  |
| **Psychiatric disorders** |  |  |  |  |  |  |
| Total ADRs | 5 (0.30) | 8 (0.15) | 37 (0.10) | 0 | 1 (0.44) | .94 |
| Fatalities | 0 | 0 | 0 | 0 | 0 | - |
| **Hepatobiliary disorder** |  |  |  |  |  |  |
| Total ADRs | 0 | 5 (0.09) | 2 (0.01) | 1 (37.40) | 1 (0.44) | - |
| Fatalities | 0 | 0 | 0 | 0 | 0 |  |
| **Skin and subcutaneous tissue disorders** |  |  |  |  |  |  |
| Total ADRs | 40 (2.40) | 18 (0.33) | 16 (0.05) | 0 | 6 (2.67) | .25 |
| Fatalities | 0 | 0 | 0 | 0 | 0 | - |
| Epidermal | 36 (2.16) | 15 (0.27) | 12 (0.03) | 0 | 6 (2.67) | .25 |

**Figure 2.**
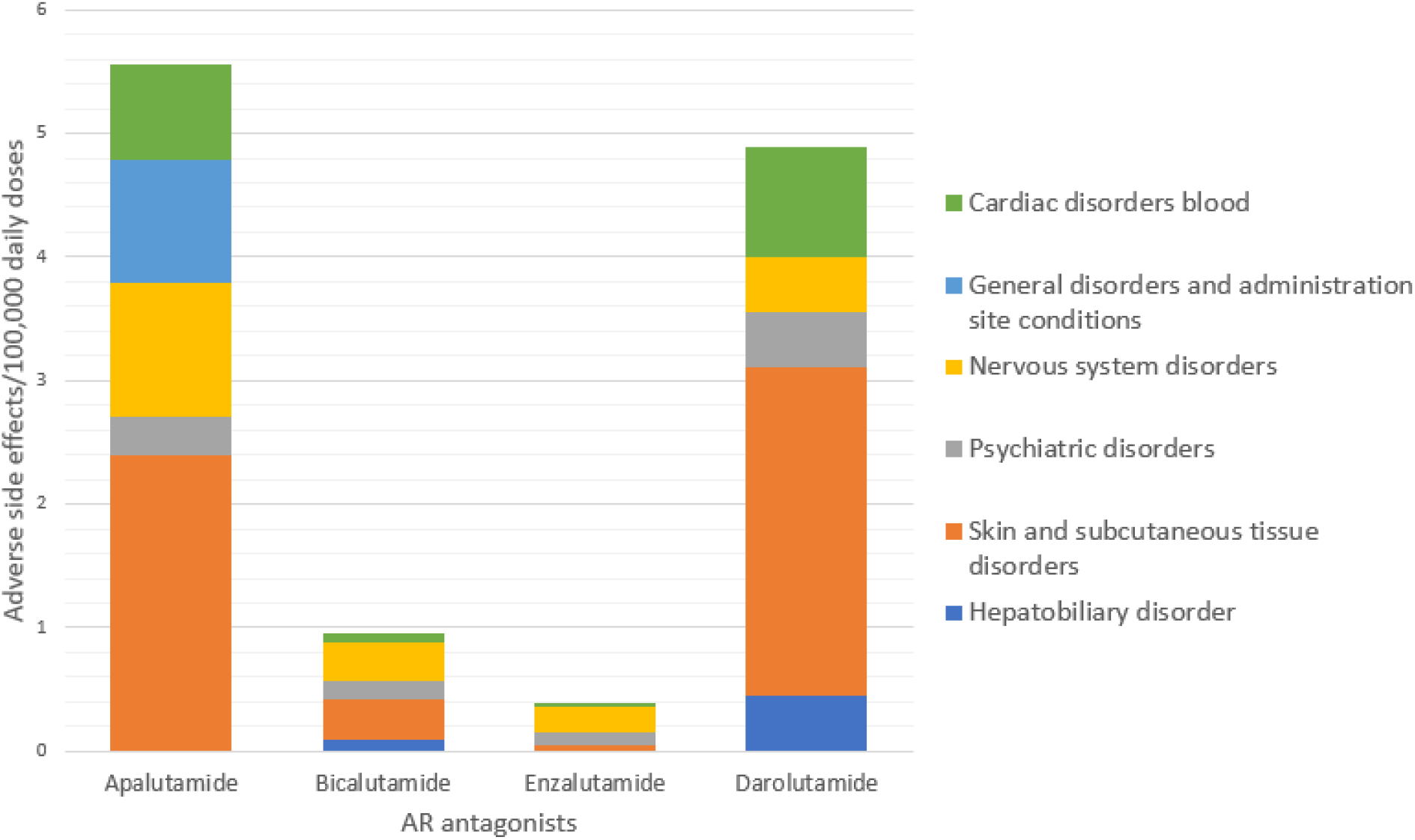
Bar chart comparison of the ADRs per 100,000 *dd* for the main SOC areas excluding flutamide (*n* = 1 ADR in these six major SOCs).

### Blood and lymphatic system disorder ADR’s and fatalities

Darolutamide and apalutamide had the highest reports of 0.44 and 0.24 ADRs per 100,000 *dd*, respectively. Patients taking darolutamide experienced suspected neutropenia (0.44/100,000 *dd*) and apalutamide experiencing suspected thrombocytopenia (0.12/100,000 *dd*).

### Cardiac disorders ADR’s and fatalities

Darolutamide had the highest suspected cardiac disorder profile (0.89/100,000 *dd*), apalutamide had the second highest ADRs/100,000 dd (0.78), along with being the only AR antagonist to have a suspected fatality in this category. The main cardiac event possessed by apalutamide patients were cardiac arrhythmias and heart failure, 0.3 and 0.36 per 100,000 *dd* respectively. Whilst enzalutamide exhibited the least cardiac events (*n* =12, 0.03 per 100,000 *dd*).

### General disorders and administration site conditions ADR’s and fatalities

General disorders and administration site conditions had the highest number of ADRs across all AR antagonists. Enzalutamide had a fatality rate of 0.8 per 100,000 *dd*. Apalutamide also experienced a high prevalence of general disorders side effects (2.04/100,000 *dd*) and fatalities (0.06/100,000 *dd*). Darolutamide followed with 1.78 ADRs/100,000 *dd*.

### Nervous system ADR’s and fatalities

Similarly, to general disorder ADR’s, apalutamide (1.08/100,000 *dd*) and darolutamide (0.44/100,000 *dd*) had the highest nervous system ADRs. Apalutamide’s main nervous system ADR was neurological disorders (0.48/100,000 *dd*).

### Psychiatric disorders ADR’s and fatalities

Darolutamide had 0.44 ADRs per 100,000 *dd*, which was the highest number of suspected events for the NSARA series. Patients on enzalutamide experienced 0.1 suspected psychiatric reactions /100,000 *dd*.

### Hepatobiliary disorders ADR’s and fatalities

The highest suspected ADRs of patients with hepatobiliary disorders were experience with flutamide (37.4 per 100,000 *dd*) but may be an artefact of the very low number of *dd* in this time period.

### Skin and subcutaneous tissue disorder ADR’s and fatalities

Similar to general disorders, skin and subcutaneous disorders had the largest number of suspected ADRs/100,000 *dd*. Darolutamide (2.67/100,000 *dd*) and apalutamide (2.4/100,000 *dd*) had the vast number of ADRs, with all darolutamide patients experiencing epidermal ADR’s and 2.16 ADR’s/100,000 *dd* for apalutamide.

### Age range of NSARA ADR reports

Apalutamide consisted of 72 patients who experienced a suspected ADR between the ages of 50-89 years old. Bicalutamide had 2 patients between 0-19 years old, with the remaining 35 patients between 60-99 years old. Enzalutamide consisted of 140 patients with side effects between 50-99 years old. Flutamide’s single report from the selected SOCs was from a male patient that was between 80-89 years old; and darolutamide having 14 patients between the ages of 60-89.

Molecular matched pair (MMP) analysis was possible between apalutamide versus enzalutamide (**Figure 1**) due to a single point variation present - gem-dimethyl versus *spiro*cyclobutane, respectively on the thiohydantoin scaffold. This led to a logarithmic difference in activity at the AR (63 vs 806 nM), and a 2.5-fold difference in hERG activity (6 vs 15.7 µM), comparable inhibition at the GABA receptor, and weak inhibition at glucocorticoid and progesterone receptors, respectively.

## Discussion

### Total ADR’s

The highest number of total ADRs reported was with enzalutamide (*n* = 1091), when standardised to the number of *dd*, flutamide (*n* = 423, 15.82 ADRs per 100,000 *dd*) and darolutamide (*n* = 35, 15.55 ADRs per 100,000 *dd*) emerged as have a higher suspected incidence rate. Flutamide’s data was excluded when comparing individual SOC ADRs per 100,000 *dd* as there was limited prescribing during the time frame of this study (3,000 *dd*) vs all other NSARAs 42,900,000 *dd*.

### Total fatalities

Enzalutamide emerged as having the highest number of fatalities (*n* = 50), when standardised to the number of *dd*, flutamide (*n* = 23, 897.5 fatalities per 100,000 *dd*) emerged as having a higher fatality rate. Darolutamide had zero reported fatalities during the time period of this study.

### Blood and lymphatic systems disorders ADR’s and fatalities

Darolutamide had a prevalence of 0.4 ADR’s/100,000 *dd* with one report of neutropenia. Apalutamide followed with 0.24 ADR’s/100,000 *dd*, with reports of thrombocytopenia. Interactions between apalutamide and CYP2C8 and CYP2C9 can cause drug-drug interactions (DDI),. Enzalutamide patients experiencing both (immune thrombocytopenia and neutropenia) again possibly due to the DDI’s with CYP2C8 and CYP2C9. Patients taking flutamide experienced no blood and lymphatic disorder ADR’s. None of the NSARAs studied were suspected of inducing a blood and lymphatic system fatality.

### Cardiac disorders ADR’s and fatalities

Darolutamide had the greatest prevalence of ADRs for cardiac disorders. The increase in cardiovascular events on darolutamide could be potentially ascribed to SERT inhibition, given its importance in the cardiovascular system.[38] Furthermore, darolutamide is a known CYP3A4 inhibitor related to potential DDIs. Prostate cancer patients are within the same age group to have cardiac co-morbidities. Cardiac medications are known CYP3A4 substrates (e.g. simvastatin/ atorvastatin [39]). Additionally, p-gp modulation causes disturbances in ion efflux which can be of concern to those taking ion channel inhibitors, such as verapamil and amiodarone. [40] P-glycoprotein inhibitors can be proton-pump inhibitors (e.g. omeprazole) and macrolides antibiotics (e.g. clarithromycin & erythromycin). The combination of these factors could have a detrimental impact on the patients’ health by increasing the risk for a cardiovascular event that is highly dependent on polypharmacy in this age group.

Apalutamide was a modest inhibitor of the human Ether-à-go-go-Related Gene (hERG) ion channel (IC_50_ = 6 µM vs C_max_ = 2.9 µM) and had the highest rate of suspected cardiac arrhythmia ADRs, 30-fold over the only other NSARA with this suspicion, enzalutamide, a significantly weaker hERG inhibitor (15.7 µM vs C_max_ = 7.7 µM). Apalutamide (0.36 ADR’s /100,000 *dd*) also interacts with CYP2C8, CYP2C9, CYP2C19 and P-gp. Inducing CYP2C19 can cause patients taking clopidogrel increase metabolism to its active metabolite, which has a half-life of approx. 30 minutes. This causes the therapeutic effect to diminish quickly after administration, making the patient at risk for an ischemic attack again dependent on polypharmacy confounders not available in these datasets.

Enzalutamide is also known to interact with CYP2C8, CYP2C9, CYP2C19, CYP3A4. Bicalutamide is a CYP3A4 inhibitor and p-gp inducer, cardiac events are potentially due to DDIs. Flutamide had no cardiac ADRs and no reported cytochrome P450 or p-gp interactions in contrast.

### General disorders and administration site conditions ADR’s and fatalities

A higher prevalence of ADR’s was observed with enzalutamide, apalutamide and darolutamide for general disorders and administration site conditions. Apalutamide’s interaction with CYP3A4, CYP2C19 and p-gp can induce DDI with other drugs that utilise their metabolic pathways. Darolutamide and bicalutamide affects CYP3A4 and p-gp, whilst enzalutamide interacts with CYP3A4. Enzalutamide had 0.08 fatalities/100,000 *dd*, followed by apalutamide (0.06).

### Nervous system ADR’s and fatalities

Effects of AR on CNS and cognitive function have been reported.[41] Apalutamide and darolutamide had 1.08 and 0.44 nervous system ADRs/100,000 *dd*, respectively. These higher reports are potentially due to their unique pharmacology. Inhibition of GABA receptor can lead to increased anxiety, stress, and fear responses and apalutamide, a GABA inhibitor (IC_50_ = 3 µM vs C_max_ = 2.9 µM) had the highest relative rate of suspected nervous system ADRs at 1.08 per 100,000 *dd*. BIn clinical settings, darolutamide did not significantly alter cerebral blood flow (CBF), consistent with its low clinical and predicted BBB penetration and subsequent low risk of CNS-related adverse events. [34] In contrast, CSF measurement in dogs for apalutamide show a 2-5% penetration which may inform patient stratification. [36]

### Psychiatric disorders ADR’s and fatalities

Darolutamide and apalutamide led with the highest number of psychiatric disorders per 100,000 *dd* (0.44 and 0.33, respectively). Darolutamide was identified as the sole NSARA to show effects at 5-HT (serotonin) receptor at < 10 µM. 5-HT has a well-known role in depression, obsessive-compulsive disorder, and social phobia and there have been reports of increased depression in patients taking AR antagonists.[42] However, low BBB penetration of darolutamide made this off-target interaction an unlikely contributor to suspected psychiatric disorders reported. Similar to the nervous system side effect profile bicalutamide and enzalutamide followed with lower numbers of suspected reports, 0.15 and 0.1 ADRs per 100,000 *dd*.

### Hepatobiliary disorders

Bicalutamide and flutamide both weakly inhibited BSEP. Chronic inhibition of BSEP can result in cholestasis due to the restriction of the bile passage. [43]

Flutamide had the largest reports at 0.44 per 100,000 *dd*, this could be due to being the only NSARA that was cleared by hepatic metabolism (approximately 50%) rather than renal clearance. Darolutamide followed with 0.44 ADR’s /100,000 *dd*. Darolutamide was the only NSARA to show effects at 5-HT (serotonin) receptor at < 10 µM and an association of inhibiting 5-HT and dysregulation of crucial components of liver function including hepatic blood flow, innervation and wound healing was noted.[44]

Apalutamide and enzalutamide did not reveal significant reports of hepatic injuries, due to possessing similar drug chemical properties. They are cleared via the kidney with a renal excretion of 65% and 71%, respectively.

### Skin and subcutaneous tissue disorder ADR’s and fatalities

NSARAs exhibited suspected skin and subcutaneous tissue disorder ADRs impacting the epidermis. Apalutamide, darolutamide and bicalutamide possessed the largest number of ADR’s/100,000 *dd* (2.40, 2.67, and 0.33, respectively). This may be due to the apalutamide containing an arylamine (**Figure 1**), which are molecular characteristics known to induce skin reactions.[45] Conversely, flutamide also contains an arylamine, but there were no reports of suspected skin or subcutaneous tissue ADRs. Bicalutamide (**Figure 1**) contains a sulfonyl-arylamine which is known to induce inflammation via IgE and IgG immune pathways.[46] Resulting in patients with a sulfonyl-arylamine allergy that are taking bicalutamide to experience epidermal side effects due to the hypersensitivity as a potential cause.

#### Limitations

Reports from the Yellow Card scheme are suspected reports, and therefore, no causal relationship must be proved before submitting a report. Therefore, reported ADRs in any registry may have no defined relationship to the pharmacology of the NSARAs. Under-reporting of ADRs is also a common issue with pharmacovigilance schemes. [47] Independent datasets were used for frequency of dispensed prescriptions and frequency of reported suspected ADRs; neither dataset enables the interpretation of comorbidity and co-prescription confounders, for example. This limits the assessment of causality and identification and adjustment for these confounding factors. Prostate cancer occurs in later stages of life where issues of medications from co-morbidities can arise.

Furthermore, the prescriber’s choice of a NSARA may be influenced by known interactions (and contraindications) with drugs co-prescribed for comorbidities, i.e., there may be a significant bias by indication (or contraindication) as these comorbidities may affect sensitivity for ADRs. For example, certain NSARAs are currently cautioned as follows enzalutamide: seizures, encephalopathy, hypersensitivity, ischemic heart disease, falls and embryo-foetal tox; darolutamide: seizures, ischemic heart disease and embryo-foetal tox; and apalutamide: seizures, ischemic heart disease, fractures, falls and embryo-foetal tox. The study cannot adjust for these confounders. There is also a NSARA dose versus risk of association with an ADR that cannot be described within these datasets.

## Conclusions

The detection of suspected ADR’s of NSARAs and potential correlations to their respected polypharmacology and physiochemical properties was identified. The highest number of ADRs were associated with enzalutamide (*n* = 1,091) and bicalutamide (*n* = 749). Enzalutamide was found to have the most off-target pharmacological interactions of the NSARAs studied (*n* = 5) including potent inhibition of GABA receptor (IC_50_ = 2.6 µM vs C_max_ = 7.7 µM) and had the most ADR reports for nervous system disorders (*n* = 72, accounting for 73% of all NSARA ADRs in this SOC). Apalutamide, the only other GABA inhibitor (IC_50_ = 3 µM vs C_max_ = 2.9 µM) had the highest relative rate of suspected nervous system ADRs at 1.08 per 100,000 dd.

Apalutamide was a modest inhibitor of the hERG ion channel (IC_50_ = 6 µM vs C_max_ = 2.9 µM) and had the highest rate of suspected cardiac arrhythmia ADRs, 30-fold over the only other NSARA with this suspicion, enzalutamide, a significantly weaker hERG inhibitor (15.7 µM vs C_max_ = 7.7 µM). Darolutamide was the only NSARA to show effects at 5-HT (serotonin) receptor at < 10 µM but did not translate to a correlation to psychiatric disorders due to low BBB penetration but may be associated with both cardiac and hepatobiliary suspected ADRs reported.

## Data Availability

All data produced in the present work are contained in the manuscript

## Acknowledgments

We thank OpenPrescribing, NHS secondary care medicines data set, EMC, FDA, ChEMBL, and MHRA Yellow Card Scheme for open access public data that underpinned this research.

